# Local retraining mitigates domain shift in sepsis prediction: Lessons from translating a neonatal model to mixed intensive care data

**DOI:** 10.64898/2026.08.18.26360666

**Authors:** Stella Champeaux, John Booth, Alexander Brown, Neil J Sebire, Ivana Drobnjak, Stuart Bowyer

**Author notes:** Corresponding author: (SB).

## Abstract

**Background:** Machine learning models leveraging electronic health records (EHRs) can support earlier detection of sepsis in intensive care units (ICUs). However, their clinical utility depends on reproducibility across institutions and patient populations. Building on a published pipeline from the Children’s Hospital of Philadelphia (CHOP), this study examines how a neonatal sepsis prediction framework performs and can be adapted to a range of intensive care environments, paediatric, cardiac, and neonatal, at Great Ormond Street Hospital (GOSH).

**Methods:** We extracted de-identified ICU EHR data from GOSH and applied feature derivation, unit harmonisation, and temporal sampling to align with the CHOP dataset used by Masino et al. (2019). Seven classifiers were first evaluated using CHOP-trained weights to characterise cross-domain behaviour and then retrained on local data to assess recoverability and site-specific adaptation. Model discrimination was summarised by AUC and F1, and learning curves were used to explore sample efficiency and bias–variance dynamics.

**Results:** Models achieved strong discrimination on the CHOP neonatal cohort but demonstrated reduced performance when transferred to the mixed GOSH ICU population, reflecting anticipated domain and population shift. Retraining on GOSH data restored discrimination (AUC range 0.69–0.86), with Gradient Boosting (AUC 0.86 vs AUC 0.87 at CHOP) and KNN (AUC 0.80 vs AUC 0.79 at CHOP) models performing comparably to their CHOP benchmarks. DeLong’s test confirmed statistically significant gains across all classifiers (p < 0.001).

**Conclusion:** ICU cohort and baseline demographic differences between CHOP and GOSH introduced domain shift that limited direct model transfer. Elements of the original preprocessing pipeline could not be reproduced, further constraining transportability. Yet, retraining on local data restored high discrimination, showing that the modelling framework remains robust when re-estimated in new settings. These results highlight local adaptation as a practical route to recover performance and support safe, generalisable deployment of clinical prediction models in mixed clinical environments.

## Introduction

### Sepsis in neonatal and paediatric intensive care

Sepsis, a dynamic and multifaceted condition, remains a leading cause of morbidity and mortality in neonatal and paediatric intensive care. Neonatal sepsis is defined as sepsis in infants under 28 days old, and is further classified as early-onset when occurring within 72 hours of birth and late-onset thereafter (1). In the neonatal population, infections occur in 6-8 per 1,000 live births and are among the leading causes of neonatal death in industrialised economies, underscoring the urgent need for accurate prediction to identify infants at elevated risk (2). Beyond the neonatal period, paediatric sepsis is defined in the International Consensus Criteria as a life-threatening syndrome of infection-associated organ dysfunction, using paediatric-specific criteria developed to address inconsistencies in earlier definitions and limitations of adult frameworks (3). Paediatric sepsis continues to impose a substantial clinical burden, with severe cases reporting mortality rates of 10–20% and sepsis contributing to more than one-third of paediatric intensive care deaths (4). Collectively, this high disease burden and evolving age-specific definitions underscore the need for predictive approaches that support reliable early recognition across diverse paediatric critical care settings.

Variability in diagnostic criteria, limited early referral to critical care and delays in administering first-line treatment complicate estimates of true sepsis incidence and hinder timely identification (6,7). The development of predictive tools is essential to efficiently identify high-risk infants. Although scoring systems such as Paediatric Early Warning Scores (PEWS) can be used to support early recognition of deterioration, these have limited specificity for sepsis and modest predictive accuracy (8). Their design prioritises bedside usability over high-resolution, data-driven inference, making them insufficient for early infection detection in intensive care environments (9). These limitations motivate the development of more precise, data-enabled predictive models capable of supporting clinical decision-making at earlier stages of illness.

### Machine learning for early sepsis prediction and the reproducibility challenge

Extensive research has demonstrated the potential of machine learning (ML) and statistical modelling to address challenges in sepsis recognition and care management (10–17). The increasing accessibility of clinical datasets, including linked EHRs, creates a promising landscape for the development of novel sepsis risk prediction models (18). In the intensive care setting, complementary work has focused on continuous physiological monitoring to anticipate deterioration before overt clinical signs. For example, models integrating heart-rate dynamics with oxygen saturation features have identified late-onset sepsis in very-low-birth-weight infants and demonstrated robust performance (AUCs > 0.82 when validated across multiple NICUs), supporting the generalisability of physiological signal–based early warning (19).

Despite this progress, reliable clinical deployment of machine learning models to different sites remains a challenge. EHRs provide the foundation for predictive modelling by capturing rich, longitudinal representations of patient care, yet they are notoriously difficult to work with, due to variations in data structure, clinical documentation practices, and coding standards across institutions create fundamental barriers to model portability and reproducibility (20). Considering these challenges, rigorous evaluation on data from outside the development environment is essential to establish trust and assess model robustness. External validation, the evaluation of a trained model’s performance on independent datasets not used during its training phase, remains the primary safeguard of generalisability. In the current UK AI landscape set by the NHS AI Lab Roadmap, this process is essential to ensure the reliability and ethical deployment of predictive models in diverse healthcare settings, cultivating trust among healthcare professionals and patients (22,23).

Regrettably, significant obstacles hinder the external evaluation of machine learning models and their seamless integration into clinical care. In a 2025 systematic review of real-time sepsis prediction models, Wang *et al.* found that fewer than **55%** of studies performed *full* external validation, and that model accuracy consistently dropped under more rigorous validation conditions (23). Often models are narrowly tailored to a single disease area or clinical department and are evaluated only on local data, limiting scalability and generalisability (24–27). The use of proprietary datasets and computationally intensive modelling methods also hinders adoption, as it requires advanced technical capabilities for successful integration (28). Moreover, integrating a model into existing electronic health record systems after development on retrospective data demands considerable investment of time and resources (29).

Together, these challenges highlight that prediction models rarely transfer seamlessly between institutions, and that static, one-off external validation is increasingly insufficient for safe deployment (30–32). Even within a single site, underlying data distributions evolve over time as patient populations, clinical workflows, and recording systems change, a phenomenon known as temporal or data drift. To achieve practical and trustworthy clinical AI, models must therefore be viewed not as static products, but as dynamic tools that require continuous local adaptation and re-evaluation (30). Approaches such as retraining on institution-specific data, recalibrating outputs to local prevalence, and aligning feature extraction pipelines with site-specific EHR structures can help bridge this gap. Researchers increasingly agree that recurrent local validation and periodic recalibration may be necessary to sustain accuracy as both institutional and temporal shifts occur (33).

### From CHOP to GOSH: testing cross-institutional generalisability

One way to examine these challenges empirically is to evaluate how an existing, well-performing model behaves when applied in a fundamentally different institutional context. A landmark study from the Children’s Hospital of Philadelphia (CHOP) demonstrated the potential for machine learning prediction of sepsis in infants hospitalised in the neonatal intensive care unit (NICU) several hours before clinical recognition, using an openly available dataset and code repository (34). In their discussion, the authors emphasised that the modelling framework was designed to be broadly applicable across institutions, noting that all features were derived from routinely collected EHR data and that the approach should be generalisable with retraining outside the NICU setting. This provides a clear motivation for evaluating how the same framework performs when applied within a different healthcare system and a more heterogeneous intensive care population.

Great Ormond Street Hospital (GOSH) is an international centre of excellence in paediatric care based in the United Kingdom. It manages approximately 76,000 children annually across 250,000 admissions and appointments (35). As an early adopter and pioneer of the intelligent hospital approach, GOSH utilises modern a modern EHR linked to a secure Trusted Research Environment (TRE), with access for approved research projects (36). GOSH therefore provides an ideal test case for validating the CHOP approach, given its role as a specialist children’s hospital with advanced EHR infrastructure. Unlike CHOP, GOSH is a specialist referral-only hospital without a labour ward, serving a highly surgical and medically complex patient population.

Definitions of intensive care units differ substantially between institutions, and patient populations within neonatal, paediatric, and cardiac intensive care units (NICU, PICU, and CICU) are shaped by local clinical practices and referral patterns (37). At GOSH, NICU primarily admits infants transferred from external hospitals for specific surgical or complex medical management. The NICU does not admit preterm babies without a need for specialist intervention. Sepsis in these patients is therefore more commonly due to postoperative complications, complications of prematurity such as necrotising enterocolitis, or late-onset, antibiotic-resistant infections, contrasting with the predominantly preterm, early-onset sepsis cohort described in the CHOP NICU (34). The CICU cares for infants and children undergoing congenital cardiac surgery, while the PICU admits a broad age range from birth to 18 years, encompassing planned postoperative cases and unplanned admissions from within the hospital or transferred from other hospitals within the regional network.

In this context, the boundaries between units are fluid: infants of the same age and with comparable physiological profiles may receive care in NICU, PICU, or CICU depending on their underlying condition, co-morbidities, and service pressures. Consequently, a “neonatal” model trained within a single ICU environment may not directly generalise across institutions where unit definitions, patient composition, and care pathways differ. In their paper, Masino et al. selected a cohort of infants under 12 months who received care exclusively in NICU, which does not map neatly onto either neonatal or paediatric sepsis definitions and excludes infants of the same age cared for in other intensive care settings. By analysing an age-matched cohort across all intensive care domains at GOSH, we aimed to provide a more realistic assessment of how the same modelling framework performs in a tertiary centre where patient flows, clinical norms, and EHR conventions span multiple related but distinct ICU environments. To our knowledge, no prior study has reproduced a published sepsis early-warning pipeline end-to-end in a new institution, nor directly compared the as-published model against an equivalent model retrained on local data. This study provides the first such evaluation within a mixed neonatal–paediatric ICU environment.

### Objectives

This study aimed to investigate the challenges of the direct transfer of a published neonatal sepsis prediction model to a specialist tertiary / quaternary paediatric centre. Our objectives were to:

- Determine the extent to which the data preparation and feature engineering methods used to derive the CHOP sepsis dataset could be replicated within the GOSH EHR environment, identifying practical and structural barriers to reproducibility.
- Assess cross-institutional generalisability by applying the CHOP-trained models to the broader GOSH ICU cohort.
- Characterise sources of domain shift by comparing demographic, clinical, and data-level features between the CHOP and GOSH cohorts.
- Assess the degree to which model performance could be restored through local retraining on GOSH data.
- Analyse the implications of these findings for the broader adoption and implementation of clinical AI models in heterogeneous healthcare environments.

We hypothesised that model parameters trained in a single institutional and clinical context are not inherently generalisable, but that the underlying feature representations and modelling framework remain transferable across domains when re-anchored to local data through retraining.

## Methods and Materials

ICU data from GOSH spanning 2019–2024 were obtained via the institution’s standardised extraction pipeline within the TRE. Records were de-identified before transfer to the study workspace, and all analyses were conducted on de-identified data in a secure NHS TRE. The dataset retained event dates (e.g. admission dates) but excluded direct identifiers (e.g., names, hospital numbers). Dates of birth were rounded to the 15th of the recorded month to maintain approximate age information while further reducing re-identification risk. In accordance with institutional policy and ethical guidance, individual consent was not required. Ethical approval was granted by the London–South East Research Ethics Committee (REC; 21/LO/0646).

### Data pre-processing

We accessed the publicly available data and full model repository from CHOP, using the original code and model specifications without modification (34). Data preprocessing followed the methodological framework described by Masino et al. (2019), with adaptations required to accommodate differences in electronic health record structure and data capture at GOSH. Core design elements, including observation windows and feature set composition, were retained to ensure conceptual alignment. However, several processing steps (e.g., cohort identification, timestamp alignment and feature summarisation) could not be replicated exactly due to differences in data resolution and availability. These adaptations were necessary to ensure data integrity and reflect local EHR conventions, consistent with the study’s aim to evaluate cross-institutional generalisability rather than strict external validation.

#### Cohort identification

All ICU-coded ward stays recorded in the GOSH EHR between 2019 and 2024 were screened. Infants were included if i) the duration of their ward stay was at least 48 hours, and ii) they were 12 months of postnatal age or younger at the time of their admission. This resulted in an eligible cohort of 2,839 unique infants. For infants with multiple ICU admissions (up to 13 per child), we selected the earliest admission to avoid mixing physiological states across distinct care episodes. Following extraction, the cohort spanned the neonatal intensive care unit (NICU), paediatric intensive care unit (PICU), cardiac intensive care unit (CICU), and the cardiac high dependency unit (CHDU).

A key methodological divergence from the CHOP study concerned the role of ‘*sepsis evaluations’* in cohort construction. Masino et al. restricted inclusion to infants who had at least one documented sepsis evaluation, a centre-specific event capturing the moment of clinical suspicion. Several categories of evaluations were then excluded, including negative evaluations (negative cultures with less than seventy-two hours of antibiotics), non-bloodstream infections, viral or fungal pathogens, and indeterminate cases such as possible contaminants or antibiotic courses between 72 and 120 hours. These evaluations were judged ineligible for modelling and these observation windows were removed; infants whose only evaluations fell into these categories were excluded entirely from the study cohort. This process produced a bacterial-sepsis–focused cohort in which both cases and controls were drawn solely from infants undergoing bacterial sepsis evaluations, resulting in a narrower and more curated population than a typical ICU census.

This evaluation-based framework could not be reproduced at GOSH. Sepsis evaluation timestamps are not included in the public CHOP release and are not consistently identifiable within the GOSH EHR, where local documentation practices differ and thresholds for initiating sepsis evaluation vary. Replicating CHOP’s inclusion and exclusion criteria would therefore require assumptions about clinician behaviour and documentation that are not transferable across institutions and could introduce systematic bias driven by local practice rather than clinical phenotype.

For these reasons, we did not apply evaluation-based inclusion or exclusion criteria at GOSH. Instead, we included all eligible ICU admissions that met the age and duration criteria, yielding a more comprehensive and unbiased representation of the paediatric critical care population at GOSH. This approach preserves the conceptual intent of the CHOP design, a cohort of infants at risk of sepsis during a period of critical illness, while avoiding reliance on documentation structures and clinical workflows that are specific to the CHOP environment. It also aligns with the study’s aim of assessing cross-institutional generalisability rather than strict replication of a single-centre inclusion rule, as this approach yielded a broader and more representative ICU cohort.

#### Case-control identification and observation windows

As sepsis evaluations could not be reproduced within the GOSH EHR, sepsis episodes were defined directly from structured microbiology and antibiotic administration data. Two case categories were created to mirror the conceptual intent of the CHOP definitions. Culture-positive sepsis was defined as isolation of a bacterial pathogen from blood culture. Clinically positive sepsis was defined as at least 120 hours of uninterrupted antibiotic therapy with the medication indication recorded as ‘Treatment of suspected sepsis’. This approach identifies episodes in which clinicians treated presumed bacterial infection in the absence of a positive culture. Although this operational definition differs from the evaluation-based rule used at CHOP, both frameworks capture clinically suspected bacterial sepsis and are conceptually aligned for the purposes of cross-institution comparison. For each sepsis episode, a 44-hour observation window was constructed spanning 48 to 4 hours before the treatment or culture timepoint, matching the temporal framing used by Masino et al. For clinically positive episodes, the indexed time was defined as the timestamp of antibiotic initiation associated with suspected sepsis, as no analogue to CHOP’s ‘sepsis evaluation time’, the time at which a clinician first suspected sepsis, was available. Using these criteria, 246 infants met culture-positive and 744 met clinically positive definitions, giving 990 sepsis episodes among 2,839 infants.

Control periods were identified using criteria adapted from Masino et al. Eligible control windows began at least 48 hours after ICU admission, occurred at least 10 days before any sepsis episode, and at least 10 days prior to death. Within each eligible control period, a single 44-hour observation window was randomly selected, consistent with the framing of the original study. In both CHOP and GOSH datasets, the modelling unit was the observation window. The CHOP cohort included multiple windows per infant, whereas in the GOSH cohort infants contributed at most one case window and one control window, reflecting differences in data structure and cohort construction. This resulted in 1,931 patients contributing control windows.

Two aspects of the original CHOP observation window methodology could not be reproduced. First, Masino et al. weighted window selection by time of day; this procedure was insufficiently described and could not be operationalised reliably. Second, their dataset allowed multiple observations per infant and used random sampling with replacement to achieve predefined case–control ratios. In our setting, resampling was not used: the GOSH cohort contained sufficient case and control windows without augmentation, and duplicating windows from the same infant would have disproportionately weighted certain clinical episodes and distorted the empirical distribution of the cohort. Each eligible observation window was therefore included once without replacement. Despite these differences, the proportions of culture-positive (8.7%) and clinically positive (26.2%) sepsis in the GOSH cohort closely matched those reported by CHOP (9% and 25%), indicating that the case-mix structure remained broadly comparable.

#### Feature extraction and harmonisation

The full set of 35 features used in this study was reproduced directly from the original CHOP model developed by Masino et al. and was therefore inherited rather than chosen independently. This includes all numerical variables, threshold-based indicators (e.g., temperature <36°C or >38°C; FiO₂ ≥40%), and binary clinical features (e.g., apnoea/bradycardia/desaturation, indwelling lines, comorbidities). Numerical features were harmonised by applying systematic unit conversions and matching the precision used in the CHOP dataset to ensure compatibility of feature scales (Table S1). One feature, the immature-to-total neutrophil (I/T) ratio, could not be derived because its constituent laboratory components were not available within the GOSH TRE.

Several binary variables describing clinical status, presence of indwelling lines, and use of advanced support therapies lacked explicit operational definitions in the published CHOP documentation (34). For reproducibility, these variables were mapped using structured diagnostic and procedural coding systems available at GOSH, including ICD-10 and SNOMED-CT terminology (Table S2). This ensured that binary features captured conceptually equivalent clinical states while relying exclusively on audit-ready, standardised codes.

This harmonisation process preserved the intent and composition of the original feature set while adapting the operational definitions to reflect the data structures available within the GOSH EHR.

#### Time-series sampling and alignment

Masino et al. did not perform time-series modelling; instead, each 44-hour case or control window was collapsed into a single feature vector, using summary statistics (mean, minimum, maximum, and last value) for each variable. Only these final aggregated features, one value per feature per observation window, were made publicly available, meaning that the underlying time-course data and exact summarisation rules could not be reconstructed.

To produce a dataset aligned in structure and intent with the published CHOP cohort, a reproducible single-value sampling method was implemented for the GOSH data. Irregular recording frequencies across features (for example, temperature measured hourly versus weight recorded intermittently) and gaps caused by inter-ward transfers led to inconsistencies in measurement timing. To address this, observations were restricted to the defined case–control windows, eligible measurements were grouped, and data were restructured so that each feature could contribute multiple time-stamped observations (Fig 1).

**Fig 1.**
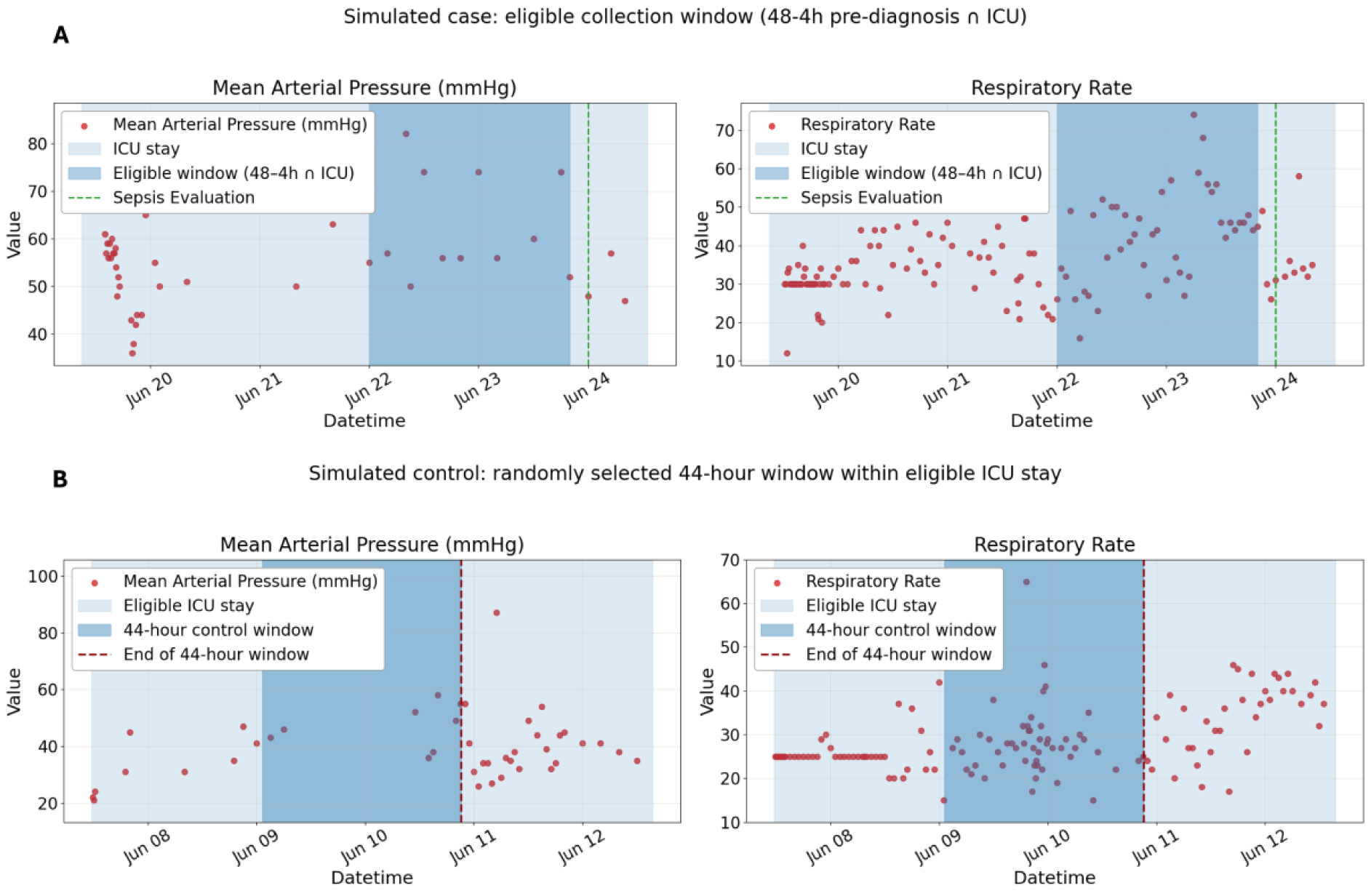
Illustration of the eligible data collection windows for sepsis cases and controls. This figure uses simulated data for illustrative purposes. Panels display Mean Arterial Pressure (MAP) and Respiratory Rate time series for simulated (A) cases and (B) controls. For cases, all measurements within the 44-hour eligible window (dark blue shading) overlapping the ICU stay and spanning 48 to 4 hours before the sepsis evaluation (green dashed line) were retained. For controls, eligible data were restricted to the ICU stay period extending up to 10 days before death or discharge, with a randomly selected 44-hour control window highlighted (dark blue shading) preceding a random control time (red dashed line). In both cohorts, multiple observations per feature frequently occurred within these windows, necessitating a single-value sampling approach to derive one representative measurement per patient for model input.

Timestamps were rounded to hourly intervals to identify each patient’s most information-rich hour; the time point containing the greatest number of concurrent features. For each feature, the value closest to this reference point was retained provided it was within the observation window, yielding a single aligned set of contemporaneous measurements per patient (Fig 2). This approach preserved the conceptual intent of capturing a representative physiological state within each observation window while maximising data availability and ensuring reproducibility within a single-row-per-patient dataset. Figures 1 and 2 use simulated data for illustrative purposes only.

**Fig 2.**
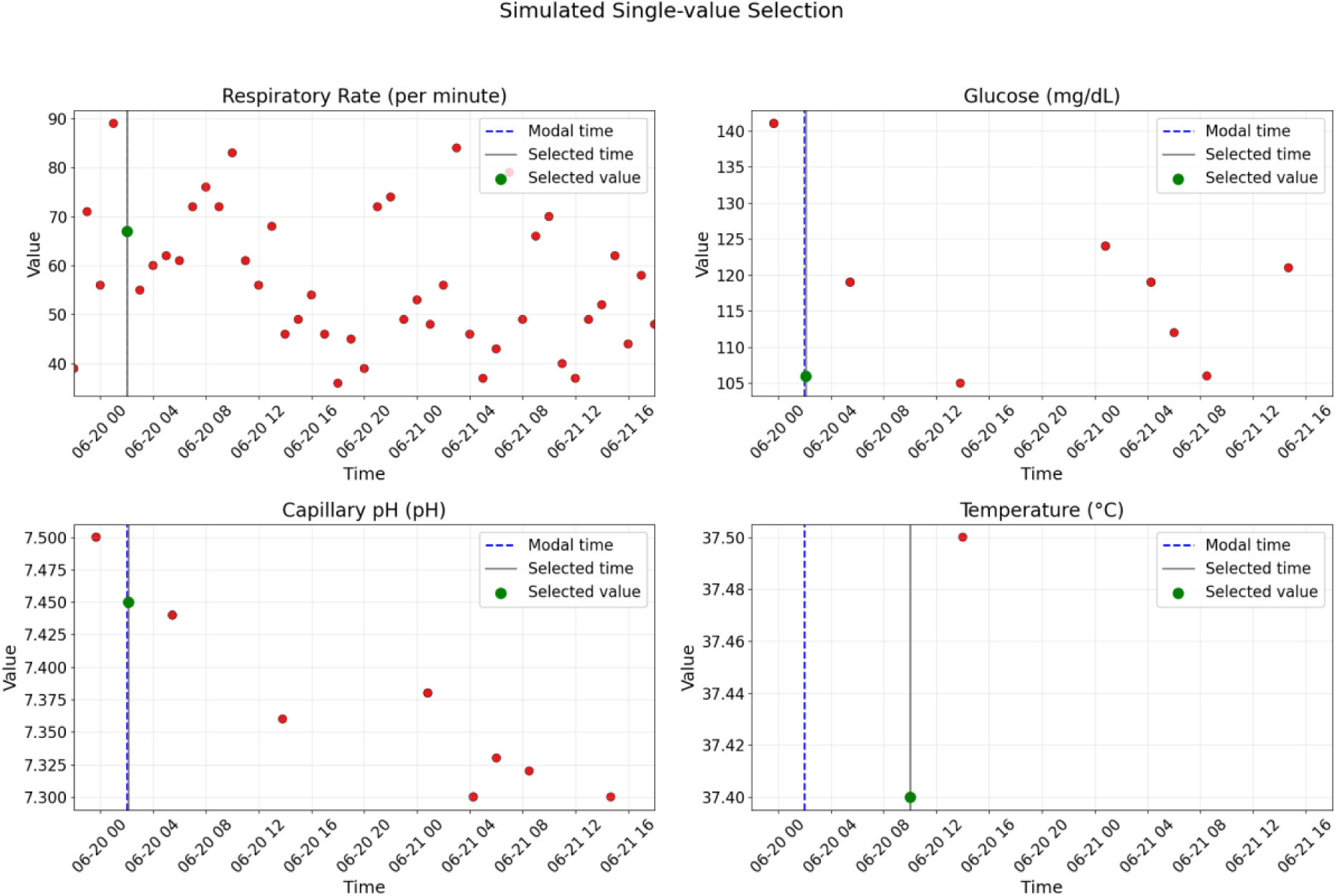
Single-value sampling via modal-time anchoring using simulated data. This figure uses simulated data for illustrative purposes. For each patient, timestamps were rounded to 1-hour bins and the modal timestamp, the hour with the highest number of distinct feature recordings, was identified. To preserve interpretability, we selected values that were temporally co-located across features, yielding an accurate snapshot of the patient’s condition. Within each feature, the snapshot value was taken as the observation closest in absolute time to this modal hour (ties resolved deterministically), yielding one value per feature per patient and enabling the pivoted matrix used for modelling. The figure illustrates this rule for four features: each panel shows all eligible measurements (red dots), the modal timestamp (blue dashed line), and the selected snapshot (green marker at its time). This modal-time anchoring handles heterogeneous sampling across features and maximises data density at selection.

#### Delta feature computation

In the original study, delta features were defined as the difference between the most recent value and the preceding 24-hour mean for heart rate, temperature, respiratory rate, and mean arterial pressure (34). Given that sampling in the GOSH dataset was irregular; to maximise data availability we adapted this by defining the delta as the difference between the snapshot value (at the modal timestamp) and the mean of all earlier measurements for that feature within the 44-hour observation window. If the modal timestamp occurred before any earlier measurements were available, the delta could not be calculated and was recorded as missing. This provides a patient-specific measure of short-term deviation from baseline at the point of greatest data density.

#### Handling of missing data and scaling

Prior to model training, we quantified missingness for each feature (Supplementary Table S3) and applied mean imputation following the approach of Masino et al.(34). No features met the CHOP-defined exclusion criteria for low availability in the GOSH cohort (>80% missing in cases or >90% in controls). Capillary pH was excluded to maintain alignment with the CHOP feature set, as this feature had removed in their pipeline due to low data availability. For both CHOP and GOSH, imputation was performed separately within each cross-validation fold: the imputer was fit on the training data only and then applied to the corresponding validation or test fold to prevent information leakage. The same fold-specific procedure was used for z-score normalisation of continuous variables. All features were imputed using the fold-specific mean except the immature-to-total neutrophil (I/T) ratio, which was imputed using the CHOP population mean as the feature could not be reliably reconstructed in the GOSH dataset. Using the CHOP reference mean ensured consistency across sites while avoiding systematic bias.

### Model development

Model development followed the methodological framework outlined by Masino et al.(34), retaining the same model architectures, feature set, and cross-validation approach to ensure comparability while re-implementing the workflow in Python using scikit-learn. This was made possible by the availability of the complete code repository and finalised dataset, which were published in full alongside the original paper.

Seven supervised classification models were trained to discriminate between sepsis-positive and sepsis-negative windows in the CHOP dataset: Logistic Regression (L2-regularised), Naïve Bayes, Support Vector Machine (SVM), K-Nearest Neighbours (KNN), Gaussian Process Classifier, Random Forest, and XGBoost. AdaBoost was excluded due to excessive training time within the TRE. SVM models used a radial basis function (RBF) kernel, with inverse-regularisation and kernel coefficient tuned over the same hyperparameter grid used by Masino et al (34). The feature space was low-dimensional (35 predictors), making all selected algorithms appropriate for this setting. While Masino *et al.* reported results separately for culture-positive and clinically suspected sepsis, in the present analysis these groups were combined into a single case cohort for all model development and evaluation.

Models were trained on the CHOP dataset using the same stratified 10-fold nested cross-validation and hyperparameter optimisation framework as the original study. Stratification ensured that each fold preserved the overall proportion of sepsis-positive and sepsis-negative windows, reducing variability in performance estimates for this imbalanced dataset. Hyperparameter tuning was conducted within each outer training fold using grid search to maximise the area under the receiver operating characteristic curve (AUC). Feature selection replicated Masino et al.’s approach by applying a mutual-information filter (SelectKBest) within each training fold to reduce overfitting. Consistent with the original implementation, cross-validation was performed at the observation-window level, as Masino et al.’s published code treats windows as independent modelling units and does not group windows by infant.

All models were implemented in Python (scikit-learn) with automated training, feature selection, and performance evaluation using a reproducible pipeline that mirrored the open-source framework published by Masino et al.

### Model transfer and local retraining

In the direct transfer experiment, the fully trained CHOP models were applied unchanged to the harmonised GOSH dataset. No model parameters were re-estimated at this stage; the transferred models retained the coefficients, tree structures, and kernel representations learned from CHOP and were applied using the same feature definitions and pre-processing steps. For models requiring feature scaling (Logistic Regression, SVM, Gaussian Process, and KNN), the original CHOP-fitted scaling parameters were applied directly to the GOSH data, ensuring that no preprocessing steps were re-estimated during direct transfer.

For the local retraining experiment, models of the same architecture were re-initialised and fitted on the GOSH dataset using the same nested cross-validation structure, mutual-information feature selection procedure, and hyperparameter search space defined in the CHOP framework. For models requiring feature scaling (Logistic Regression and SVM), standardisation was applied within each fold using parameters estimated from the training split only, mirroring the CHOP implementation. The SVM used an RBF kernel with the same grid-search ranges for C and γ as reported in the original study, and the

Gaussian Process Classifier was implemented using the scikit-learn RBF kernel with default hyperparameters, consistent with Masino et al. and computationally feasible given the dataset size. Masino et al. did not specify whether windows from the same infant were kept within the same cross-validation fold; their published code uses window-level stratified folds (34). In this study, patient-level grouping was implemented to prevent information leakage arising from multiple correlated windows contributed by the same infant, which is standard best practice when repeated observations are present.

All analyses were conducted in Python 3.11 (scikit-learn) within the GOSH Trusted Research Environment. Fold-level AUCs and predicted class-probabilities were exported for side-by-side comparison of transferred and retrained models. Differences in paired AUC estimates were assessed using DeLong’s test, with p < 0.001 considered statistically significant.

Following Masino et al.,(34) we also generated learning curves to characterise how model performance changed with increasing training set size. Examining these curves across both CHOP and GOSH datasets allowed us to assess whether observed differences in discrimination were attributable to model structure or to differences in cohort size and heterogeneity.

## Results

### Cohort characterisation and dataset comparison

To contextualise the cross-site performance patterns, we evaluated differences in the underlying data landscape between CHOP and GOSH. Supplementary Figure 1 shows the relative distribution of eligible ICU admissions across differing ICU environments at GOSH. CICU accounted for the largest proportion of patients (43.6 %), followed by PICU (25.7 %), NICU (18.4 %), and CHDU (12.2 %), which is a specialised cardiac high dependency unit.

Continuous feature distributions (Fig 3 and 4) revealed systematic baseline shifts, with GOSH patients being heavier and of higher postnatal age and gestational age at birth consistent with the lack of inborn preterm children and the inclusion of a mixed intensive care population spanning NICU, PICU, and CICU settings, while CHOP included a larger proportion of very preterm infants. Binary feature comparisons (Fig 5) showed higher prevalence of surgical and congenital comorbidities and greater use of invasive interventions in GOSH, contrasted with higher necrotising enterocolitis rates at CHOP. Baseline demographic summaries (Table 1) confirmed these distinctions, but ethnicity and race categories provide descriptive comparison only and are not directly equivalent across datasets. Overall, the GOSH cohort represents a heavier, older, and more surgically complex population than the original NICU-only CHOP cohort.

**Fig 3.**
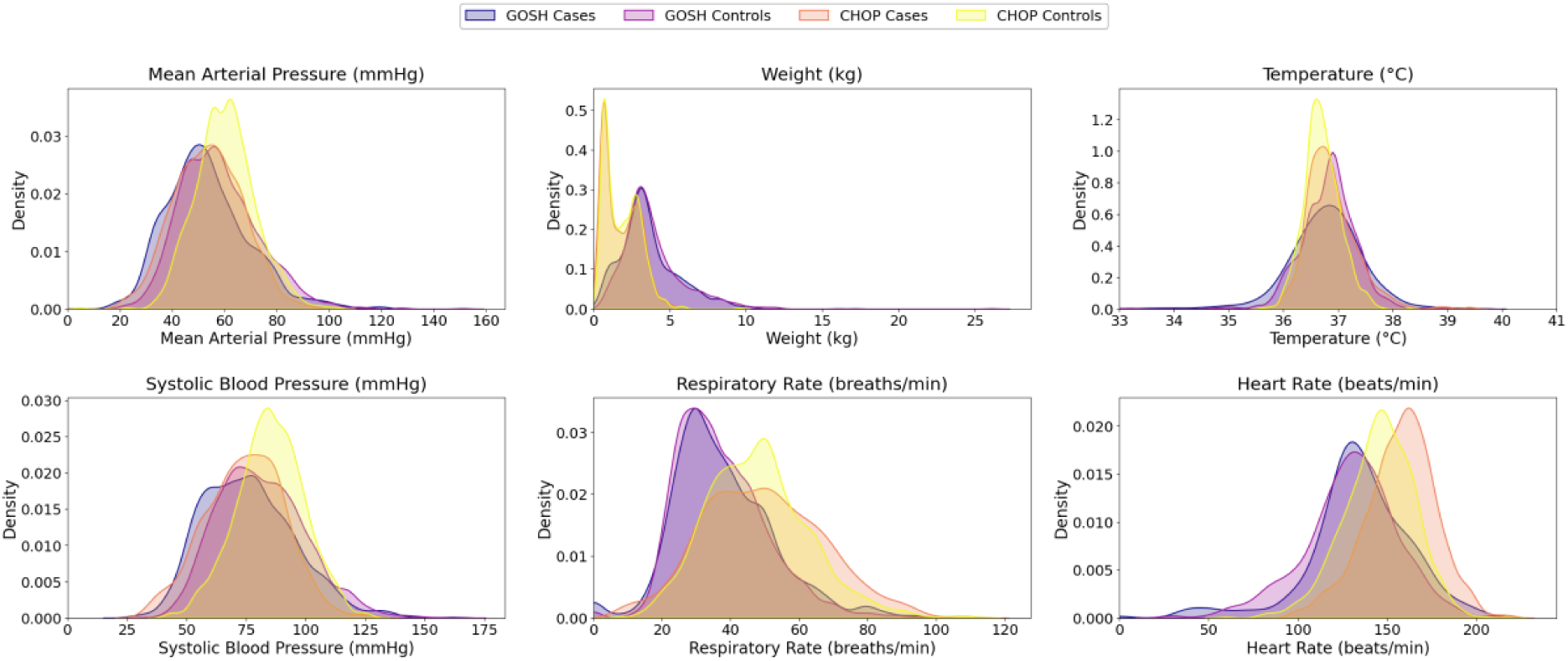
Density overlays of continuous variable features: CHOP vs GOSH cohorts. Distributions of six physiologic features included in the original Masino et al. sepsis model, shown here for GOSH and CHOP cases and controls. Compared with CHOP, GOSH patients were typically heavier. Greater divergence between CHOP cases and controls is observed for several vital signs, including heart rate, respiratory rate, and mean arterial pressure.

**Fig 4.**
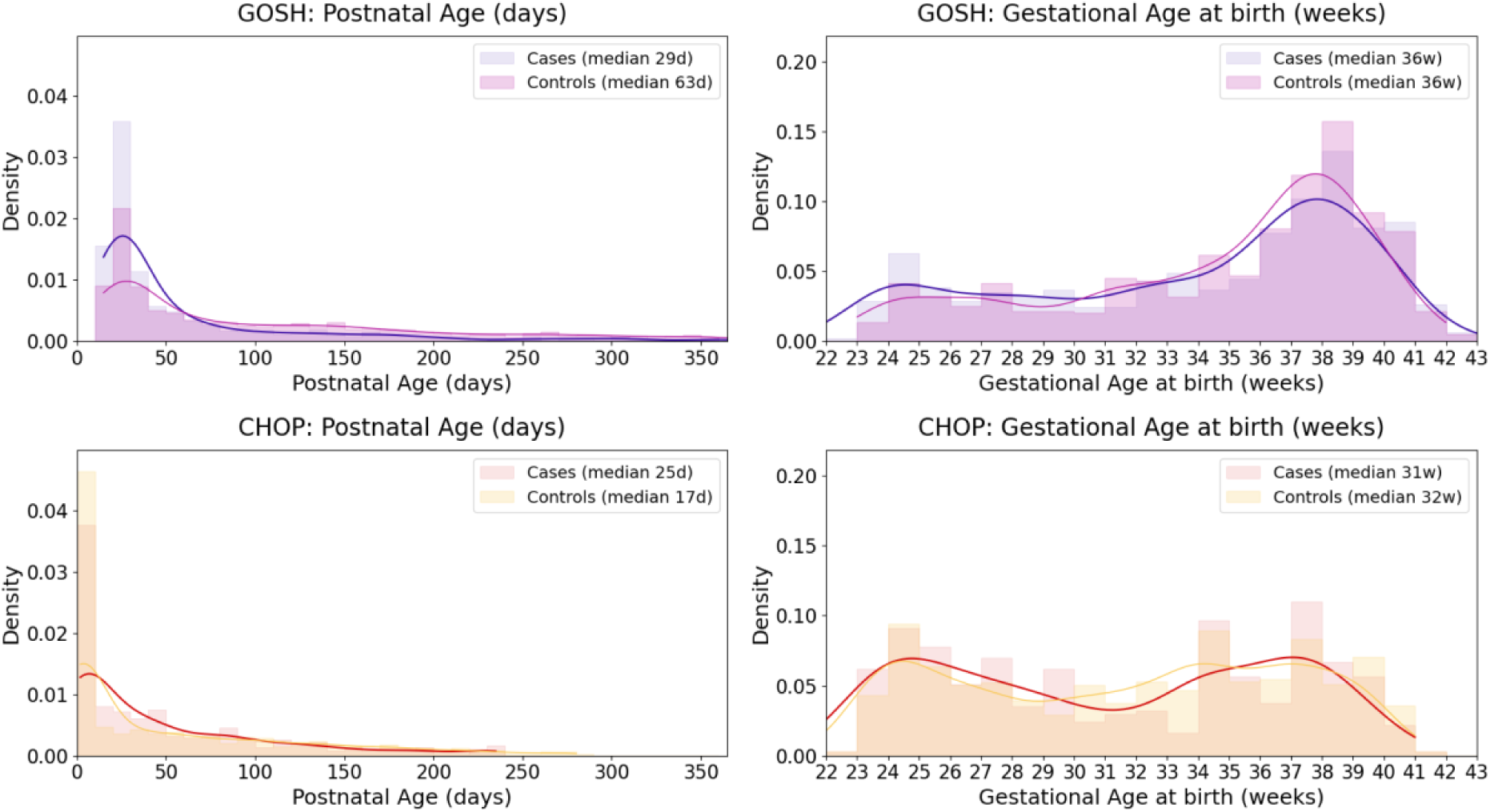
Age distribution of cases and controls in the GOSH and CHOP cohorts. Distributions of postnatal age and **ge**stational age at birth for cases and controls in the GOSH (Panel A) and CHOP (Panel B) cohorts. These features are shown because they were selected as predictive variables in the original sepsis model by Masino et al. (2019). GOSH patients were generally older postnatally and more often born at later gestational ages compared with CHOP patients. At GOSH, postnatal ages show an apparent cutoff below 14 days; this reflects an artefact of the de-identification process in which dates of birth were rounded to the 15th of the month, affecting infants within their first two weeks of life.

**Fig 5.**
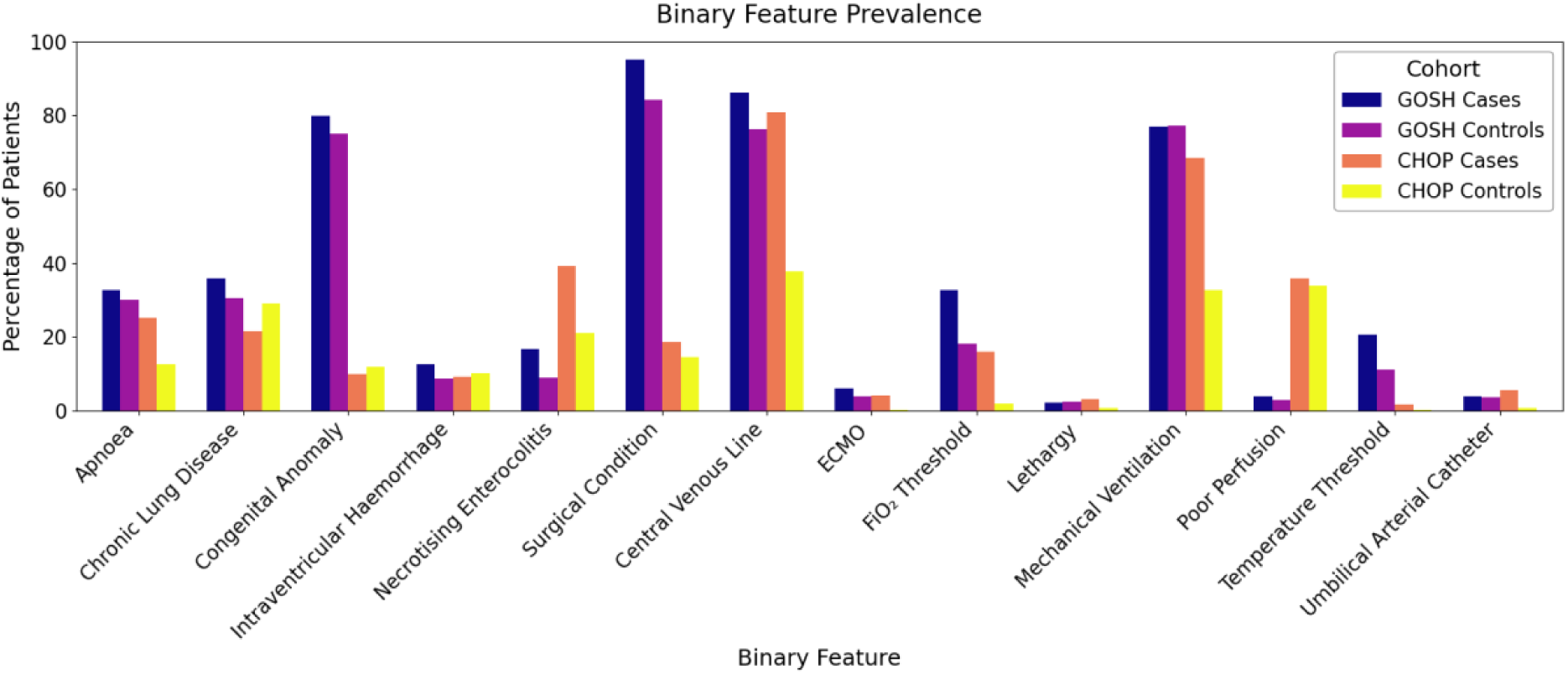
Prevalence of binary clinical features across CHOP and GOSH mixed-ICU sepsis cohorts. All binary variables shown here were inherited directly from the original CHOP model developed by Masino et al. and were therefore reconstructed in the GOSH dataset using ICD-10 and SNOMED-CT mappings (operational definitions in Table S2). Threshold-based indicators were defined according to the CHOP specifications: temperature threshold was set to 1 if temperature was <36°C or >38°C and 0 otherwise, and FiO₂ threshold was set to 1 if FiO₂ ≥40% and 0 otherwise. GOSH patients demonstrated higher rates of congenital and surgical conditions, consistent with the hospital’s role as a tertiary and quaternary referral centre and showed greater use of intensive interventions such as central venous lines and mechanical ventilation. In contrast, CHOP cohorts exhibited higher prevalence of necrotising enterocolitis and poor perfusion.

**Table 1.**
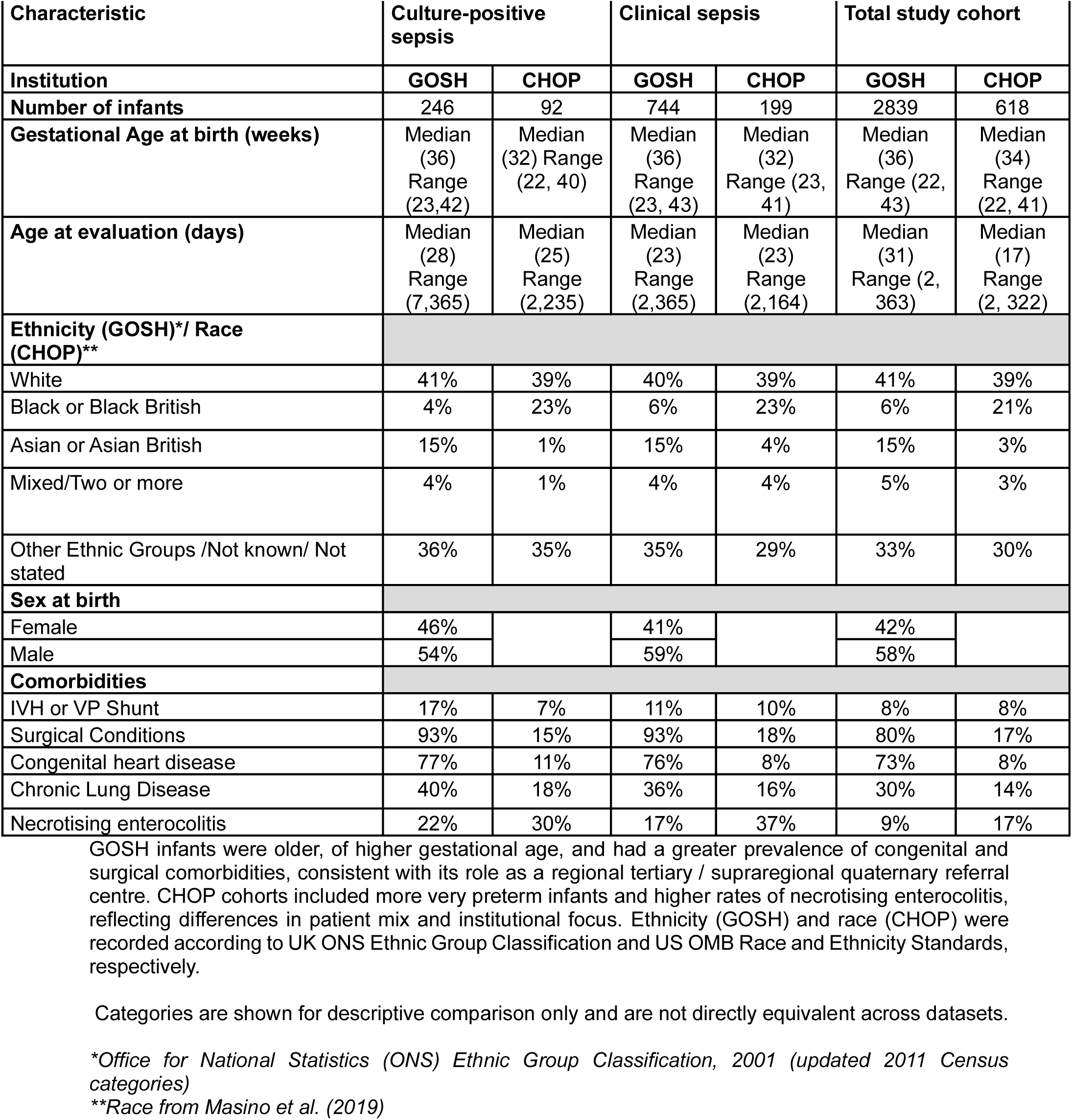
Baseline demographics of infants across GOSH and CHOP mixed-ICU cohorts.

Examination of feature completeness (Supplementary Fig 2) indicated that while routine physiological measures were well captured across both datasets, laboratory features were more complete in GOSH, and time-series variables such as heart rate were unexpectedly sparse, reflecting differences in data extraction pipelines. Mean imputation reduces the discriminative value of continuous variables with high missingness (e.g., heart rate and gestational age), and this effect is more pronounced at GOSH, where several physiological features are sparsely documented. Finally, feature importance analysis (Fig 6) demonstrated that variables with strong predictive weight in CHOP, such as necrotising enterocolitis (NEC), contributed less within GOSH, whereas indicators of surgical complexity were more salient locally. NEC is particularly sensitive to local diagnostic and documentation practices, and the marked difference in its predictive value likely reflects variation in coding as well as possible epidemiological differences between institutions. Collectively, these findings reflect the substantial demographic and structural differences that underpin the domain shift between these two paediatric centres and help explain the observed generalisation gap between cohorts.

**Fig 6.**
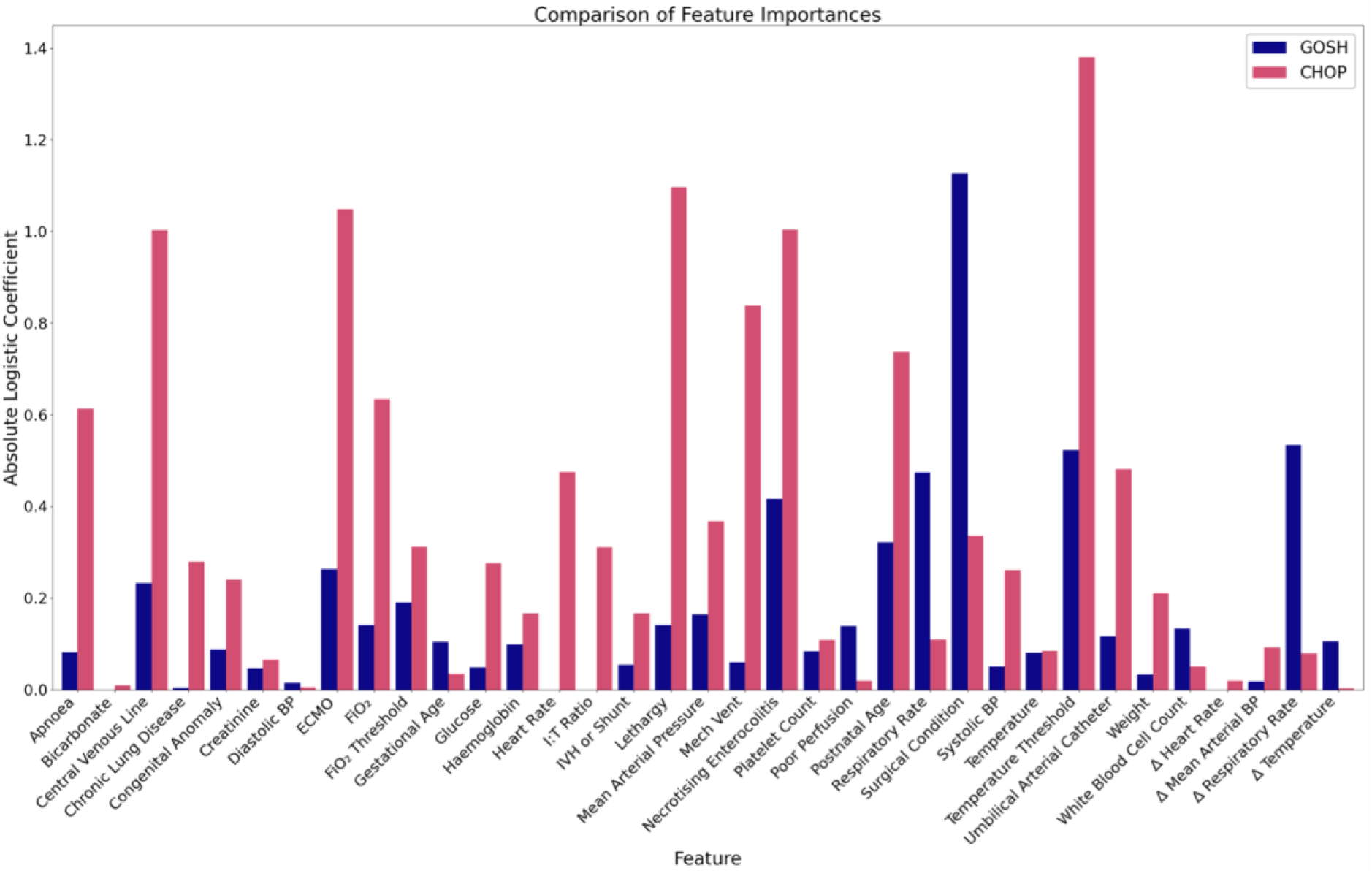
Comparative analysis of feature importance in the logistic regression model for CHOP and GOSH data. Bars show the absolute logistic coefficients for each feature in models trained on CHOP (pink) and GOSH (blue). Higher values indicate greater influence on sepsis prediction. Differences reflect variation in feature availability and distribution between sites.

### Model performance across sites

The development (training) AUCs obtained by rerunning the original CHOP code on the CHOP dataset exactly matched the values reported in Masino et al. for the combined clinically suspected and culture-positive sepsis cohorts, confirming that the published CHOP implementation is internally reproducible when executed on the original dataset. Table 2 summarises model discrimination across internal evaluation at CHOP, direct transfer to GOSH, and local adaptation following retraining on GOSH data. Figure 7 visualises these trajectories, highlighting the characteristic drop in AUC on direct transfer and the heterogeneous recovery after local adaptation. Across models, discrimination was strong in development at CHOP, with AUCs ranging from 0.79 (Gaussian Process and KNN) to 0.87 (Gradient Boosting). There was anticipated attenuation on direct transfer at GOSH, where AUCs fell (AUC: 0.62 to 0.65). Retraining models locally on GOSH data substantially restored discrimination, with AUCs approaching or exceeding those observed in the original CHOP setting. For instance, Gradient Boosting achieved the highest overall performance across all three settings (AUC: 0.87 in development, AUC: 0.63 in direct transfer and AUC: 0.86 in local adaptation), confirming its robustness and adaptability. Ensemble and kernel-based methods such as Random Forests, Support Vector Machines, and Gaussian Processes displayed similar recovery after local adaptation, whereas simpler linear and probabilistic models (Logistic Regression and Naïve Bayes) remained modestly lower. Collectively, these findings demonstrate that while a fall in performance on direct model transfer is expected, local retraining effectively restores predictive discrimination across institutions.

**Table 2.** Area under receiver operating characteristics.

| Model | Internal Performance<br>(CHOP) | Direct Transfer<br>(CHOP → GOSH) | Local Adaptation<br>(GOSH) |
| --- | --- | --- | --- |
| Gradient Boosting | 0.87 [0.83, 0.93] | 0.63 [0.61, 0.65] | 0.86 [0.82, 0.90] |
| Gaussian Process | 0.79 [0.72, 0.86] | 0.65 [0.65, 0.66] | 0.78 [0.75, 0.82] |
| KNN | 0.79 [0.73, 0.85] | 0.64 [0.63, 0.65] | 0.80 [0.76, 0.84] |
| Logistic Regression | 0.85 [0.80, 0.91] | 0.62 [0.62, 0.64] | 0.71 [0.69, 0.76] |
| Naïve Bayes | 0.84 [0.80, 0.89] | 0.65 [0.65, 0.65] | 0.69 [0.62, 0.73] |
| Random Forest | 0.86 [0.82, 0.94] | 0.62 [0.60, 0.64] | 0.78 [0.73, 0.83] |
| SVM | 0.86 [0.79, 0.92] | 0.65 [0.58, 0.66] | 0.79 [0.77, 0.85] |

AUC values summarise model discrimination across three settings: internal evaluation within CHOP (cross-validation), direct model transfer from CHOP to GOSH without retraining, and local model adaptation following retraining on GOSH data. Values are mean AUCs over 10 nested cross-validation iterations; brackets show the fold-wise range. Higher AUC values indicate stronger discrimination, with 0.5 representing chance-level performance and 1.0 a perfect classifier.

**Fig 7.**
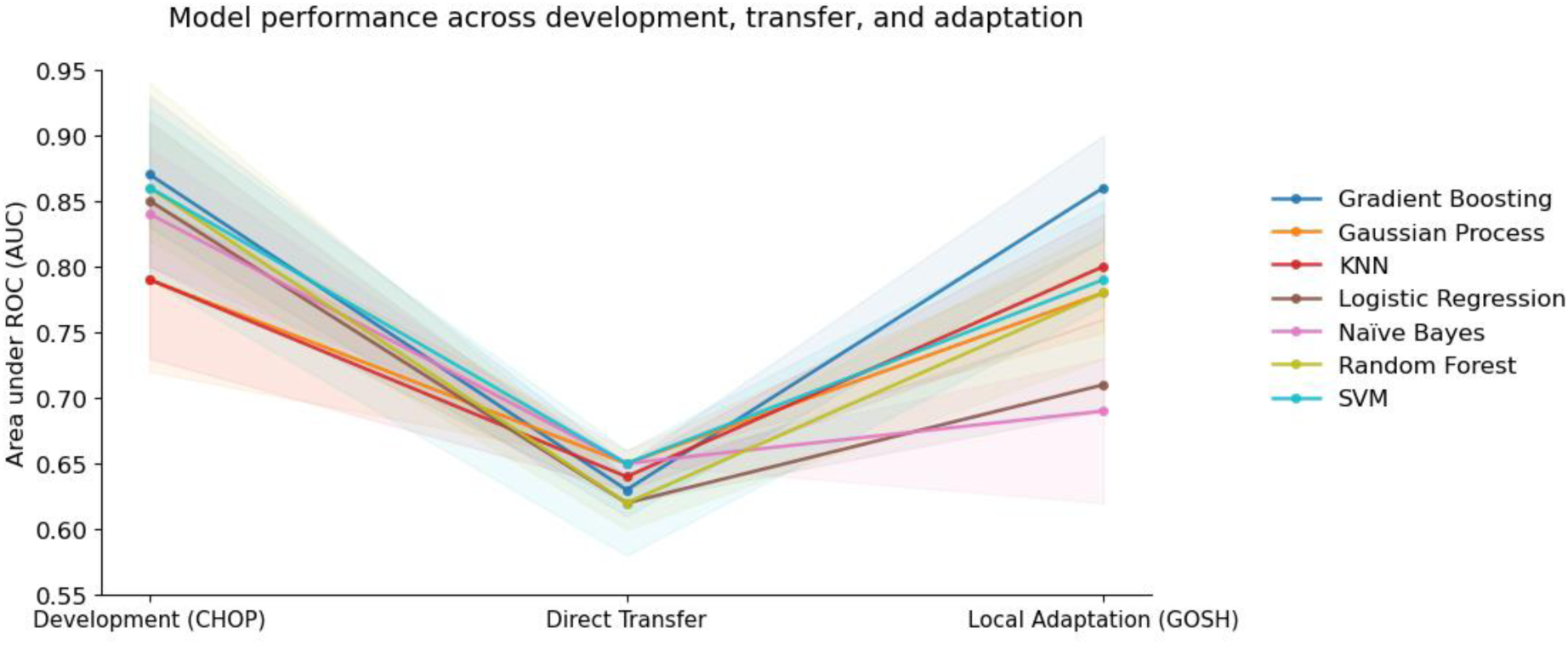
Model performance across development, cross-site transfer, and local adaptation. Visualisation of Area under the ROC curves (AUC) for seven machine-learning models evaluated in three settings: model development on CHOP data, direct cross-site transfer to GOSH without retraining, and local adaptation following retraining on GOSH. Lines show mean AUC across 10 nested cross-validation iterations, and shaded regions denote the fold-wise minimum and maximum values. All models exhibit a marked decline in discrimination when transferred across institutions, followed by heterogeneous recovery after local adaptation.

Supplementary Figure S3 shows ROC curves for Logistic Regression, SVM, and Gradient Boosting across the three evaluation settings. Each panel displays the ROC curves from the 10 outer cross-validation folds together with the median ROC curve. The clustering of fold-wise curves toward the upper left during CHOP development, their shift toward the diagonal at GOSH under direct transfer, and their upward recovery after local retraining mirror the AUC patterns summarised in Table 2.

### Precision-recall performance

F₁ scores, reflecting the balance between precision and recall, followed the same cross-site trend as AUC; high during CHOP development, reduced on direct transfer at GOSH, and restored after local retraining (Table 3). Gradient Boosting and Random Forest achieved the highest F₁ values throughout (CHOP 0.61–0.65; GOSH retrained 0.61–0.68), consistent with their ability to model nonlinear interactions and remain robust under class imbalance. Gaussian Process and KNN performed consistently lower, reflecting their limited extrapolation capacity and sensitivity to local data density. Naïve Bayes displayed variable results, with marginal apparent improvement on direct transfer but lower scores after retraining, likely reflecting instability from its strong feature independence assumptions.

**Table 3.** F1 scores.

| Model | Internal Performance<br>(CHOP) | Direct Transfer<br>(CHOP → GOSH) | Local Adaptation<br>(GOSH) |
| --- | --- | --- | --- |
| Gradient Boosting | 0.61 [0.48, 0.70] | 0.49 [0.47, 0.51] | 0.68 [0.59, 0.73] |
| Gaussian Process | 0.48 [0.40, 0.50] | 0.39 [0.37, 0.41] | 0.59 [0.49, 0.67] |
| KNN | 0.47 [0.39, 0.57] | 0.38 [0.36, 0.42] | 0.59 [0.47, 0.63] |
| Logistic Regression | 0.64 [0.55, 0.74] | 0.51 [0.50, 0.51] | 0.56 [0.50, 0.62] |
| Naïve Bayes | 0.53 [0.37, 0.71] | 0.55 [0.52, 0.56] | 0.49 [0.48, 0.50] |
| Random Forest | 0.64 [0.50, 0.69] | 0.51 [0.50, 0.53] | 0.61 [0.55, 0.67] |
| SVM | 0.65 [0.57, 0.73] | 0.53 [0.52, 0.53] | 0.61 [0.53, 0.66] |

Values represent mean $F_1$ scores averaged over 10 nested cross-validation iterations, with brackets showing the fold-wise range. Higher $F_1$ values indicate a stronger precision–recall balance at the chosen classification threshold.

Learning curves for Gradient Boosting, SVM, and Logistic Regression are shown in Supplementary Figure S4. As expected, training F1 decreases and validation F1 increases with larger training sets, reflecting reduced variance and improved generalisation as more data become available. Models trained on CHOP reach stable performance at relatively small sample sizes, consistent with CHOP being a more homogeneous neonatal cohort with less variability in physiological patterns. In contrast, GOSH models require substantially more data before convergence and reach a lower plateau, reflecting the greater heterogeneity and complexity of the mixed ICU population. The training–validation gap observed at smaller sample sizes therefore reflects population heterogeneity rather than model instability, and the eventual convergence indicates that performance in each setting is driven primarily by underlying cohort structure.

### Statistical evaluation of model retraining

To formally assess whether the observed improvements in discrimination following local retraining were statistically significant, DeLong’s test for correlated ROC curves was applied to paired AUC estimates obtained from the same cross-validation folds under the direct-transfer and retraining conditions. Retraining on the GOSH cohort improved AUCs for all classifiers, with absolute gains ranging from +0.04 (Naïve Bayes) to +0.23 (Gradient Boosting). Across all models, DeLong’s test rejected the null hypothesis of equal AUCs between the transferred and retrained models (e.g., Gradient Boosting: Z = −7.0, p < 0.001), indicating statistically significant recovery of discrimination after local adaptation (Supplementary Table S4). No adjustment for multiple comparisons was applied because each comparison reflects an independent model-specific evaluation rather than repeated testing of a single hypothesis.

## Discussion

To our knowledge, this study provides the first end-to-end reimplementation and external evaluation of a published neonatal sepsis early-warning pipeline in a different hospital, and the first direct comparison of the as-published model against an equivalent model retrained on local data. Our findings demonstrate that CHOP-trained models transferred poorly to the mixed GOSH ICU cohort, but their discrimination was substantially restored once retrained on local data. The observed recovery indicates that single-environment machine learning pipelines can be successfully adapted to mixed-ICU environments once refitted to local data. This behaviour makes the mechanism of failure, and the mechanism of recovery, explicit: the original parameters encode CHOP-specific data-generation processes, whereas retraining re-estimates those parameters under GOSH’s structure, restoring alignment between the model and its operating environment. In this sense, domain shift is not simply an observed performance drop, but a methodological signal pointing to the necessity of local parameter re-estimation.

The cross-institutional differences observed between CHOP and GOSH highlight how institutional context shapes the data signal available to machine learning models. As shown in Figures 5, 6, and 7, the GOSH cohort comprised older, heavier and predominantly term infants and exhibited a higher prevalence of surgical and congenital conditions. This reflects the hospital’s role as a tertiary and quaternary referral centre and results in a more complex, predominantly surgical case mix with fewer preterm infants compared with CHOP. These demographic contrasts have direct implications for model generalisability, as the risk profiles, physiological norms, and disease dynamics of preterm neonates differ substantially from those of older or post-surgical infants.

At CHOP, many infants present directly from labour wards and are at risk of early-onset sepsis driven by perinatal factors such as maternal infection, chorioamnionitis, or premature rupture of membranes (38). In contrast, a large proportion of infants admitted to specialist surgical environments at GOSH (especially the CICU) typically enter care for planned interventions, often required due to congenital abnormalities, rather than acute infectious presentations. Where sepsis does occur, it more typically arises post-operatively or after prolonged hospitalisation, with distinct physiological trajectories and antimicrobial susceptibilities. Our dataset does include cases where early onset sepsis co-occurs with surgical pathology, as well as admission due to a need for surgical management in the context of infection (for example, necrotising enterocolitis). However, the distribution of sepsis phenotypes in our dataset differs substantially from the largely early-onset pattern captured at CHOP, limiting model generalisability unless parameters are re-estimated on the local population.

These clinical and demographic differences manifest directly in feature–outcome relationships, underscoring the need for local adaptation. In addition to demographic differences, several binary clinical features could not be defined identically across institutions. CHOP’s definitions for variables such as NEC, respiratory compromise, and lethargy were not fully specified in the public release, and equivalent documentation structures were not available in the GOSH EHR. Accordingly, these variables were mapped using locally auditable ICD-10 and SNOMED-CT codes, introducing some definition shift that may influence their predictive value. Feature importance analysis in Figure 9 revealed that predictors with high discriminative value at CHOP, such as necrotising enterocolitis, extracorporeal membrane oxygenation (ECMO) and lethargy, were less relevant within GOSH, whereas indicators of surgical complexity and cardiorespiratory support were more informative. Since the feature selection process was inherited from the CHOP pipeline, it prioritised variables most predictive in a neonatal population, leading to reduced compatibility in a mixed ICU environment. Local retraining reweighted these relationships, aligning model coefficients with the dominant clinical patterns in the GOSH data, and likely explains the recovery in AUCs. Together, these shifts in feature relevance show that sepsis prediction models trained in a neonatal setting cannot generalise reliably to a mixed-ICU population without local retraining to realign the model with site-specific clinical patterns.

GOSH provided a uniquely challenging and data-rich environment for assessing the transportability of the Masino et al. framework. As a national specialist centre with complex surgical, cardiac and mixed-ICU case-mix, GOSH represents exactly the type of heterogeneous population that tests whether a neonatal sepsis model can generalise beyond its original clinical setting. The large cohort, availability of structured EHR data, and access to the original open-source CHOP code enabled a faithful re-implementation of the modelling pipeline and a rigorous assessment of model behaviour under domain shift. Conducting all analyses within a modern Trusted Research Environment ensured secure, reproducible handling of clinical data. Although evaluated at a single external centre, the factors driving domain shift in our study (differences in patient mix, documentation practices, EHR structure, and service configuration) are common across many hospitals. The demonstration that performance can be restored through routine retraining therefore has direct relevance for other institutions seeking to operationalise externally developed clinical AI models.

Several limitations inherent to retrospective EHR data constrained exact reproduction of the original workflow and influence interpretation of the findings. Differences in metadata availability prevented replication of CHOP’s ‘sepsis-evaluation’ construct, including their exclusion of contaminant blood cultures, viral or fungal pathogens, and indeterminate evaluations. As a result, the GOSH cohort reflects a broader and more heterogeneous real-world ICU population than the bacterial-sepsis–focused CHOP cohort. In addition, the exact reproduction of the CHOP pipeline was constrained by differences in data availability and documentation structure. Several binary clinical features (e.g., NEC, respiratory compromise, lethargy) could not be defined identically using GOSH records, as CHOP’s operational definitions were not fully specified in the public release. These variables were therefore mapped using ICD-10 and SNOMED-CT codes, introducing unavoidable definition shift. One CHOP predictor (the immature-to-total neutrophil ratio) could not be derived at all and was imputed to preserve feature dimensionality, illustrating broader challenges in cross-site data harmonisation. Irregular sampling in the GOSH dataset further required adapting CHOP’s 24-hour delta calculation. Mean imputation, retained for methodological consistency, does not capture informative missingness, and computational constraints within the TRE limited evaluation of more demanding algorithms. These factors underscore the need for standardised data capture and infrastructure to support reproducible machine-learning validation across institutions.

### Why models fail to travel and the role of local retraining

Machine learning models trained within one hospital frequently perform poorly when deployed elsewhere, despite using identical algorithms and seemingly comparable features (21,23,39). This pattern has been observed across many clinical domains and stems from the fact that prediction models encode not just statistical relationships but also the full data-generation pipeline of the environment in which they were trained, such as its extraction logic, documentation practices, cohort definitions and clinical workflows (40,41). When any of these components differ between the development site and the deployment site, even well performing models can fail to generalise (39,42). Understanding the reasons why models struggle to travel is therefore essential for designing approaches that can recover performance in new settings. Our findings illuminate key contributors to failed transportability and demonstrate how local retraining can function as an effective corrective.

A first challenge concerns differences in the underlying data architecture and preprocessing workflows across institutions. In reproducing the CHOP pipeline within the GOSH environment, several components could be implemented faithfully, but others could not be recreated due to variation in local guidelines, data organisation, metadata availability and documentation styles, constraints that are typical of retrospective EHR research and remain a persistent barrier to reproducibility (43). Even with open code, differences in data models and extraction conventions mean that implementation choices at one institution rarely translate directly to another (44). The absence of common data standards therefore continues to limit the transferability of machine learning workflows across hospitals (45). Developing shared frameworks for preparing and reporting EHR data for machine learning would greatly improve reproducibility and comparability. While NHS initiatives such as reproducible analytical pipelines and Trusted Research Environments aim to improve data quality and transparency (46,47), data sharing is still limited by interoperability and privacy constraints (48–50). Federated learning offers a promising future direction, but its success will depend on greater harmonisation of EHR structures across NHS trusts (51). These challenges highlight that even perfect conceptual reproducibility cannot compensate for divergent data infrastructures.

A second contributor to failed transportability is the mismatch between the population on which a model is trained and the population in which it is deployed. Intensive care units are organised very differently across institutions, and labels such as neonatal, paediatric, or cardiac intensive care describe local service configurations rather than fixed clinical entities (37). As anticipated, models trained on CHOP data failed to generalise effectively to a mixed-ICU environment at GOSH. At CHOP, the model was trained within a single neonatal intensive care unit predominantly caring for preterm infants with early or late onset neonatal sepsis (34). In contrast, the GOSH cohort spans neonatal, paediatric, and cardiac intensive care and includes term surgical infants, children with congenital heart disease, acutely deteriorating infants and interhospital transfers. These differences substantially reshape both the distribution and the clinical interpretation of key predictors such as ventilation, vital signs, and laboratory trends, limiting the transportability of model parameters learned in a population dominated by preterm neonates. Hence, population heterogeneity represents a fundamental limit on cross-institutional model performance.

In our case, transportability is further challenged by differences in how sepsis is labelled and documented across institutions. There is no universally accepted operational definition of sepsis for retrospective research (52), and Masino et al relied on local clinical suspicion when constructing their labels (34). While CHOP’s framework yielded high internal performance by modelling the local decision-making process, it renders the model brittle when moved to a new environment where workflows differ. At GOSH we adapted this framework using culture positivity and prolonged antibiotic treatment, but definitional practices are shaped by local antimicrobial stewardship policies, thresholds for empirical therapy and documentation behaviour (37), introducing an additional layer of label shift. As a result, the retrospective label ‘sepsis positive’ in EHR-derived datasets often reflects local clinical behaviour as much as underlying infection, even when operationalised through apparently objective events (e.g., antibiotic administration, abnormal temperature, blood culture sampling) (52). Hence, the inability to replicate this label at GOSH is not merely a data limitation, but a finding in itself: models anchored to subjective workflow markers (e.g., ’evaluation ordered’) learn to predict clinician behaviour rather than patient physiology. Retraining recovered performance across all classifiers, even though sepsis definitions and documentation differed between centres. This shows that the modelling framework can be adapted reliably through local retraining, even when labels do not align perfectly across hospitals.

Despite these challenges, our results show that these limitations are not insurmountable. Retraining the same model architectures on local GOSH data substantially restored discrimination across all classifiers, in some cases exceeding the performance originally achieved at CHOP. To our knowledge, this is the first demonstration of recovering performance by locally retraining a fully reimplemented neonatal sepsis early-warning pipeline outside its development environment. This indicates that while model parameters learned in one setting do not travel well, the underlying modelling framework can perform effectively when recalibrated to local data. From an implementation perspective, this suggests that when patient populations or documentation practices differ across institutions, or when full reproducibility is not possible, site-specific retraining offers a practical way to recover performance while respecting local variation.

Future work should focus on practical strategies to improve cross-institutional adaptation and make sepsis prediction tools more robust to variation in patient populations and EHR structure. One avenue is to initialise models with parameters learned elsewhere and fine-tune them on local data, enabling smoother adaptation than full retraining. Improving the quality and consistency of data used for modelling will also be essential: standardising physiological measurements, laboratory reporting, and clinical event documentation across NHS centres would reduce information loss during preprocessing and support more reliable feature construction. Approaches that treat missingness as informative, rather than filling gaps with simple averages, may better capture real clinical trajectories. Finally, evaluating the same modelling framework across multiple NHS paediatric centres would clarify how institutional practices shape performance and help establish shared guidelines for updating models as local data evolve.

This study addressed the challenges of translating a published neonatal sepsis prediction model to a mixed intensive care population in a different healthcare system. We provide the first end-to-end external reproduction and cross-institution evaluation of a neonatal sepsis early-warning pipeline outside its development environment. Models trained on the CHOP cohort did not generalise to GOSH, reflecting substantial differences in patient mix, clinical practice, data structure, and the reproducibility limits of the original preprocessing pipeline. By dissecting demographic, physiological, and data-quality differences, we identified clear sources of domain shift that explain the loss of performance after transfer. Crucially, retraining the same model architectures on local GOSH data restored discrimination across all classifiers, demonstrating that although learned parameters are institution-specific, the modelling framework itself remains transferable once recalibrated to local distributions. More broadly, our results show that with site-specific retraining, clinical prediction models can be transferred across institutions without loss of performance, providing a practical roadmap for centres seeking to operationalise externally developed models.

## Data Availability

The CHOP dataset and associated model code used in this study are publicly available through the repository accompanying the original study by Masino et al. The GOSH data used in this study are derived from routinely collected clinical data held within the GOSH Clinical Informatics Data Store and are not publicly available owing to patient confidentiality and information-governance requirements. Access to these data may be available to approved researchers subject to the appropriate application, governance and institutional approvals from Great Ormond Street Hospital for Children NHS Foundation Trust. Aggregate and derived results supporting the findings of this study are provided within the manuscript and its Supporting Information.

https://github.com/chop-dbhi/sepsis_01

## Acknowledgments

The authors thank Professor Mart Peters for clinical expertise in neonatal sepsis, Lucy Orr Ewing and Nirav Shah (Coalition for Health in AI) for feedback on clinical AI governance, and Erwann Le Lannou for methodological guidance.

## Supporting Information

**S1 Table**. **Unit conversions applied to align GOSH data with CHOP dataset.** Clinical features extracted from GOSH EHR required systematic unit conversion to ensure compatibility with the CHOP dataset.

**S2 Table. Coding definitions for comorbidities, clinical assessment features, and interventions.** SNOMED-CT codes were used to define nursing assessments of clinical status, while ICD-10 codes were applied to comorbidities, surgical conditions, indwelling lines & support. This coding approach ensured consistent operationalisation of binary features across the GOSH dataset for external validation.

**S3 Table. Proportion of missing data across features in GOSH and CHOP cohorts.** This table summarises the proportion of missing values for each clinical feature across case and control groups in the GOSH and CHOP datasets. Data completeness varied substantially between cohorts.

**S4 Table. Statistical comparison of AUC performance before and after model retraining using DeLong’s test.** A negative Z-score indicates higher AUCs following retraining, while the p-value denotes the significance of this difference (significance threshold p = 0.05). All classifiers demonstrated statistically significant gains in AUC on the local GOSH cohort after retraining compared with directly transferred CHOP models.

**S1 Figure.**
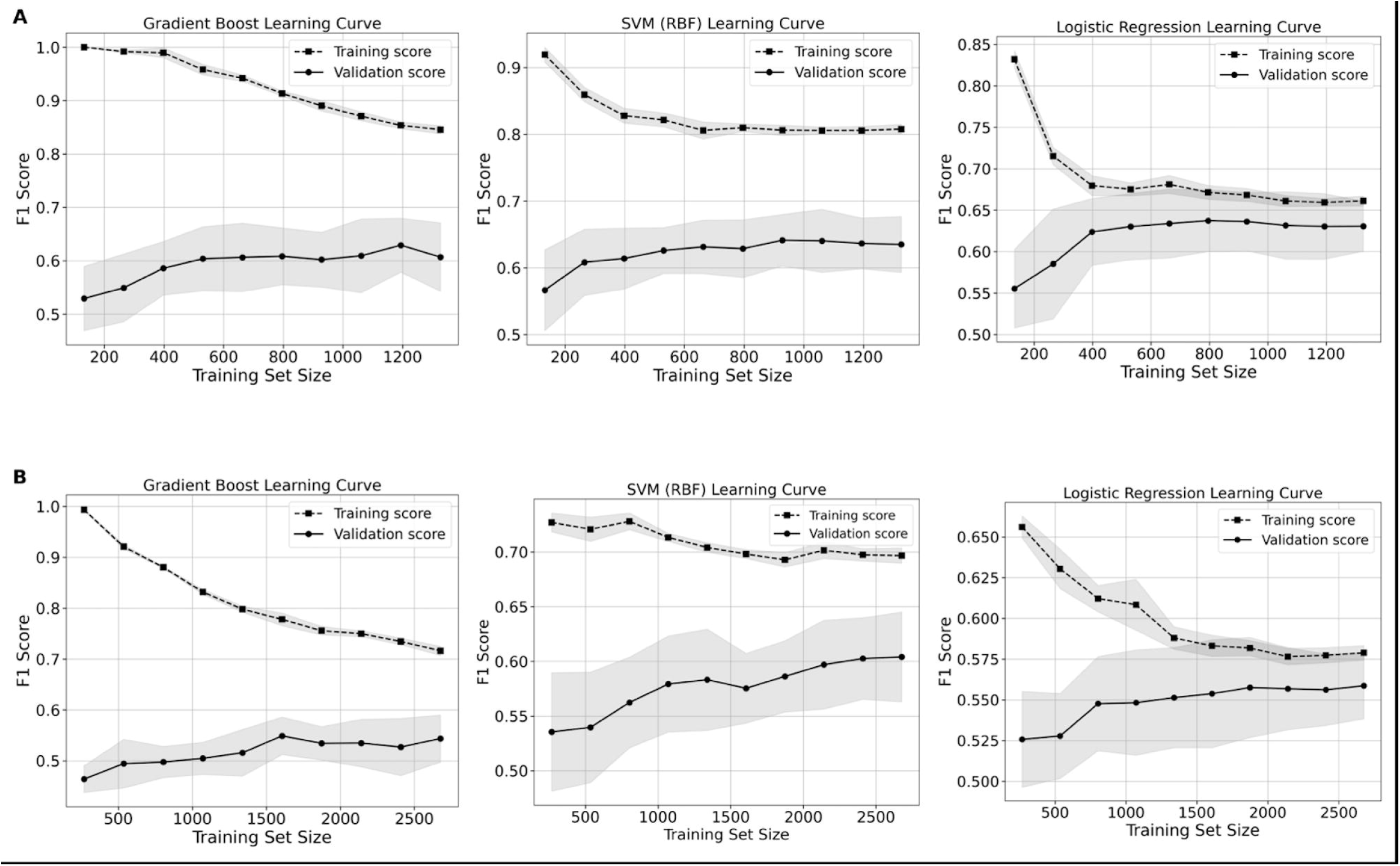
Distribution of ICU admissions by ward type following age restriction. Admissions were distributed across cardiac (CICU 43.6 %), paediatric (PICU 25.7 %), neonatal (NICU 18.4 %), and the CHDU (12.2 %), reflecting the hospital’s predominance of surgical and cardiac cases.

**S2 Figure.**
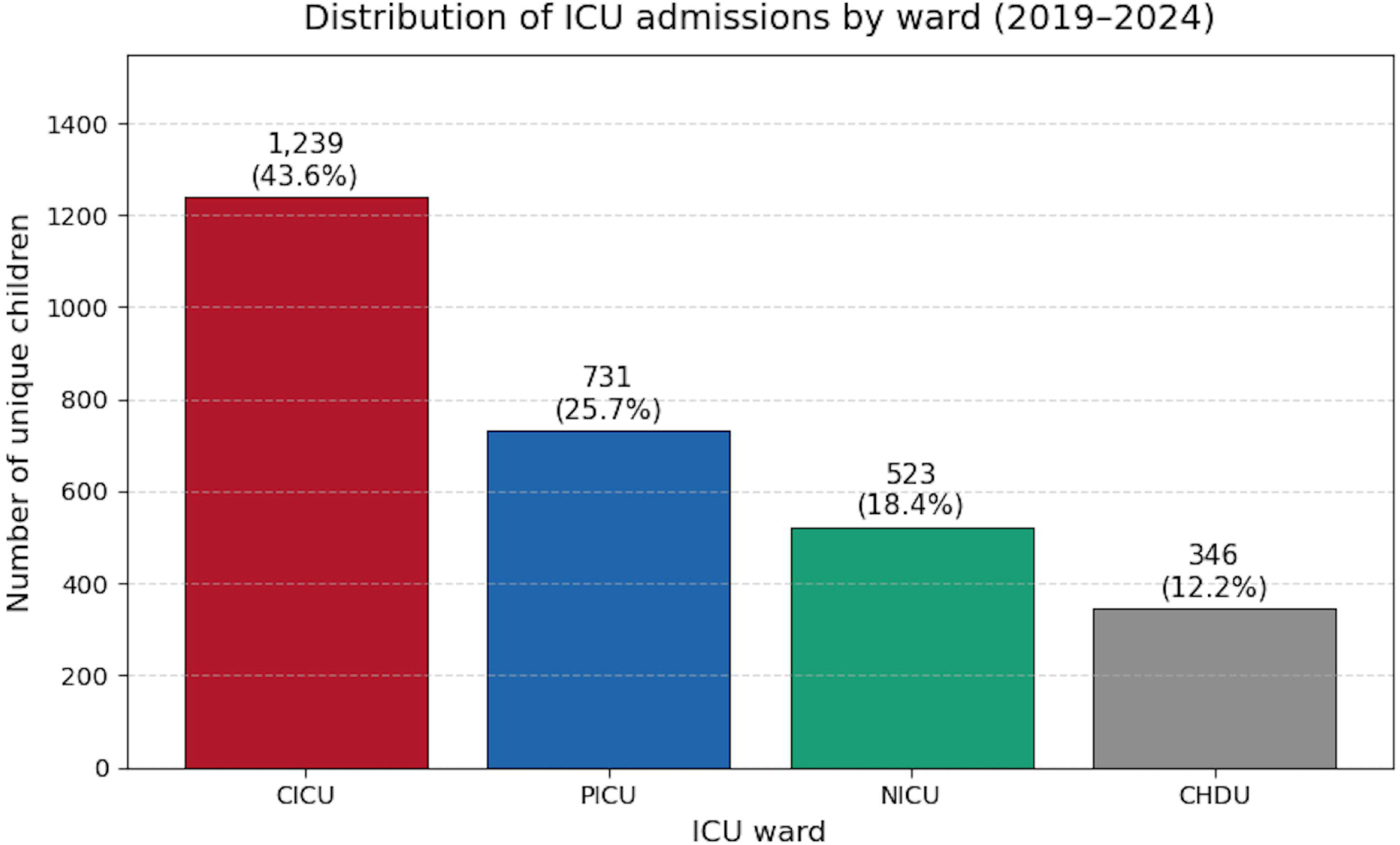
Feature availability plots in the GOSH & CHOP dataset for continuous variable features. Feature availability across GOSH and CHOP cohorts. Routine ICU measurements such as respiratory rate, temperature, blood pressure, FiO₂, and weight were consistently available across both sites. In contrast, laboratory-derived features (e.g., WBC, haemoglobin, capillary pH, creatinine) were more complete in GOSH than CHOP. Notably, heart rate availability was unexpectedly low in the GOSH dataset compared with CHOP, reflecting differences in how physiological data were recorded or extracted. These discrepancies highlight systematic site-level variation in feature completeness, which poses a significant challenge for cross-site model validation and generalisability.

**S3 Figure.**
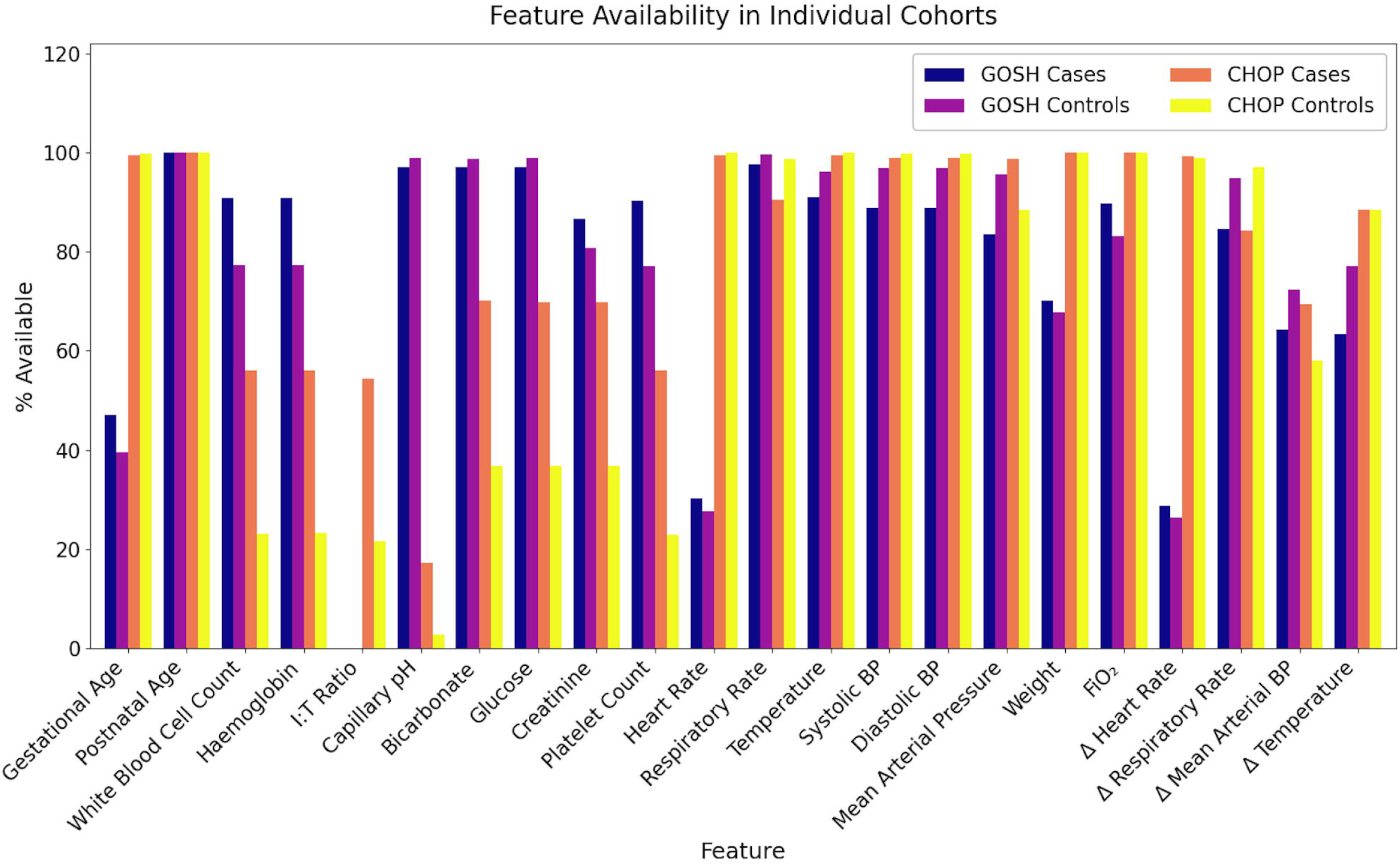
Receiver operating characteristic (ROC) curves for the Support Vector Machine (SVM), Gradient Boosting and Logistic regression models in training, direct transfer and local adaptation. Receiver operating characteristic (ROC) curves for Logistic Regression (A), Support Vector Machine (B), and Gradient Boosting (C) across internal evaluation at CHOP (top), direct transfer to GOSH (middle), and local retraining on GOSH data (bottom). Each panel shows the ROC curves from the ten outer cross-validation folds (grey) and the median ROC curve (black). Curves illustrate the anticipated decline in discrimination under direct model transfer and subsequent recovery following local adaptation.

**S4.**
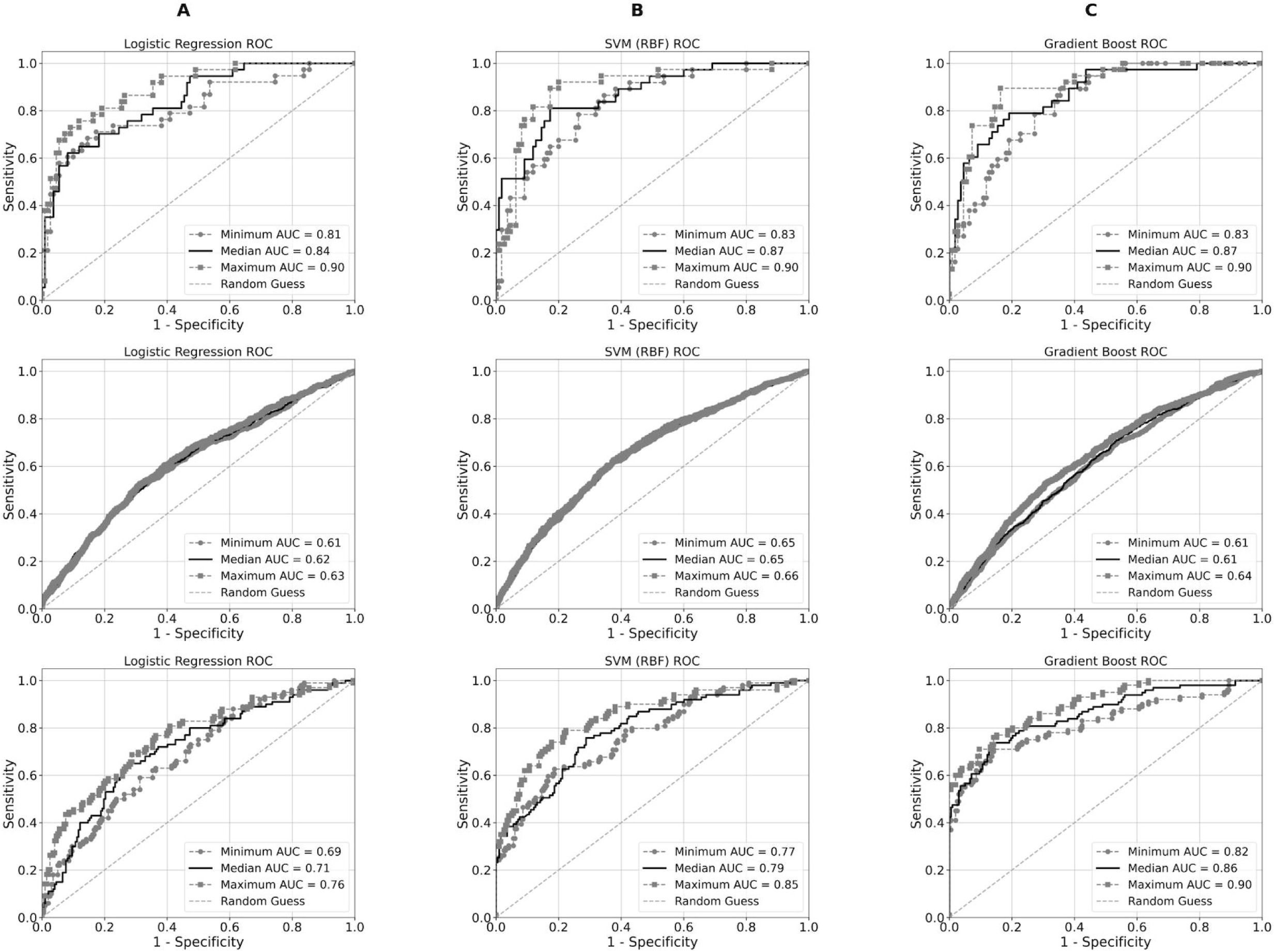
Learning curves for CHOP development and GOSH retraining. Learning curves for Gradient Boosting, SVM (RBF), and Logistic Regression during (A) CHOP development and (B) GOSH retraining. Training and validation F1 scores are shown across increasing training-set sizes, with shaded min–max envelopes. Panel A converges earlier and to a higher plateau because the CHOP neonatal cohort is more internally homogeneous. Panel B converges later and at a lower plateau despite a larger sample, reflecting the greater clinical and physiological heterogeneity of the mixed ICU population at GOSH. The expected pattern of decreasing training F1 and stabilising validation F1 with increasing sample size is observed across all models.

