## Supplementary tables 1-4 for "Local retraining mitigates domain shift in sepsis prediction: Lessons from translating a neonatal model to mixed intensive care data"

**S1 Table. Unit conversions applied to align GOSH data with CHOP dataset.**

| Feature | Original unit (GOSH) | Converted unit (CHOP) |
|---|---|---|
| Temperature | °C | °F |
| Weight | g | kg |
| Creatinine | µmol/L | mg/dL |
| Haemoglobin | g/L | g/dL |
| Glucose | mmol/L | mg/dL |

Clinical features extracted from GOSH EHR required systematic unit conversion to ensure compatibility with the CHOP dataset.

**S2 Table. Coding definitions for comorbidities, clinical assessment features, and interventions.**

| Feature Group | Feature Name | Coded Definition |
|---|---|---|
| **SNOWMED CT** | | |
| *Clinical assessments* | Apnoea, bradycardia & other desaturation events | '1023001','3415004','13094009','38593000','301795004','48867003','248583008','271825005','373895009','449171008','1179627006' |
| | Lethargy | '271795006','89458003' |
| | Poor perfusion | '309585006', '816082000' |
| **ICD 10** | | |
| *Comorbidities* | Chronic lung disease | 'Q334', 'Q322', 'P27', 'P280', 'P281', 'P282', 'P285', 'P288', 'P289' |
| | Congenital heart disease | 'Q20', 'Q21', 'Q22', 'Q23', 'Q893', 'I518' |
| | Intraventricular haemorrhage & ventriculo-peritoneal shunt | ‘P102', 'P520', 'P521', 'P522', 'P523', 'I615', 'T850' |
| | Necrotising enterocolitis | 'P77', 'A047' |
| | Other surgical conditions* | String in ‘diag_name’ contains ‘operation’ |
| *Indwelling Lines & Support* | Central venous line | 'L91','PRO85', 'ANE147','IMG90006', 'IMG155301', 'IMG90000','IMG8025', 'IMG90172', 'IMG90035', 'IMG15009', 'IMG15008', 'IMG90076' |
| | Umbilical artery catheter | 'PRO151', 'O152', 'T243', 'LAB2382' |
| | Excorporeal membrane oxygenation cannula | 'X581 |

SNOMED-CT codes were used to define nursing assessments of clinical status, while ICD-10 codes were applied to comorbidities, surgical conditions, indwelling lines & support. This coding approach ensured consistent operationalisation of binary features across the GOSH dataset for external validation.

**S3 Table. Proportion of missing data across features in GOSH and CHOP cohorts**

| Feature | GOSH Culture Positive (N = 246 patients) | GOSH Clinically Positive (N = 744 patients) | GOSH Controls (N = 1931 patients) | CHOP Culture Positive (N = 92 patients) | CHOP Clinically Positive (N=199 patients) | CHOP Controls (N = 618 patients) |
|---|---|---|---|---|---|---|
| Gestational age | 43% | 56% | 60% | 0 | <1% | <1% |
| Postnatal age | 0 | 0 | 0 | 0 | 0 | 0 |
| White blood cell count | 9% | 9% | 23% | 45% | 43% | 74% |
| Haemoglobin | 9% | 9% | 23% | 45% | 43% | 74% |
| Platelet count | 9% | 9% | 23% | 46% | 43% | 74% |
| I/T ratio | 100% | 100% | 100% | 48% | 45% | 75% |
| Capillary pH | 3% | 3% | 2% | 84% | 82% | 95% |
| Bicarbonate | 3% | 3% | 2% | 33% | 33% | 56% |
| Glucose | 3% | 3% | 2% | 33% | 29% | 56% |
| Creatinine | 15% | 13% | 19% | 33% | 29% | 56% |
| Respiratory rate | 1% | 3% | <1% | 11% | 9% | 2% |
| Temperature | 6% | 10% | 4% | <1% | <1% | 0 |
| Heart rate | 78% | 68% | 73% | <1% | <1% | 0 |
| Systolic blood pressure | 6% | 12% | 3% | <1% | 1% | 0 |
| Diastolic blood pressure | 6% | 12% | 3% | <1% | 1% | 0 |
| Mean arterial pressure | 11% | 18% | 2% | 2% | 1% | 9% |
| Weight | 29% | 30% | 32% | 0 | 0 | 0 |
| Fraction inspired Oxygen (FiO2) | 6% | 12% | 0 | 0 | 0 | <1% |

This table summarises the proportion of missing values for each clinical feature across case and control groups in the GOSH and CHOP datasets. Data completeness varied substantially between cohorts. In GOSH, gaps were most notable for gestational age (43–60%), weight (29–32%), and heart rate (68–78%), whereas most laboratory values were relatively complete (<15% missing). By contrast, the CHOP dataset showed high missingness for several laboratory features, particularly white blood cell count, haemoglobin, platelet count (43–74%), capillary pH (82–95%), and I/T ratio (45–75%). Physiological monitoring features were generally more complete in CHOP than GOSH. These discrepancies reflect structural differences in data capture between institutions and emphasise the need for harmonisation strategies before model training.

**S4 Table. Statistical comparison of AUC performance before and after model retraining using DeLong's test.**

| **Model** | **Z score** | **P-value**<br>(Significance level p = 0.05) |
|---|---|---|
| Gradient Boosting | -7.0 | < 0.001 |
| Gaussian Process | -8.7 | < 0.001 |
| KNN | -10.5 | < 0.001 |
| Logistic regression | -8.4 | < 0.001 |
| Naïve Bayes | -9.7 | < 0.001 |
| Random Forest | -7.4 | < 0.001 |
| SVM | -10.3 | < 0.001 |

A negative Z-score indicates higher AUCs following retraining, while the p-value denotes the significance of this difference (significance threshold p = 0.05). All classifiers demonstrated statistically significant gains in AUC on the local GOSH cohort after retraining compared with directly transferred CHOP models.
